# Dihydroartemisinin for Treatment of Polycystic Ovary Syndrome: study protocol for a multi-centre placebo-controlled Randomized Clinical Trial

**DOI:** 10.1101/2025.11.25.25341030

**Authors:** Liangshan Mu, Yanting Shen, Guiquan Wang, Changqin Liu, Haiyan Yang, Wentao Li, Ben Willem Mol, Xiaoying Li, Xi Dong, Jingjing Jiang

## Abstract

**Introduction:** Polycystic ovary syndrome (PCOS) is the most common endocrine disorder that affects reproductive-aged individuals. Our previous single-arm pilot trial showed that dihydroartemisinin (DHA) normalized menstrual cycles, ameliorated hyperandrogenemia, and reduced antral follicles in individuals with PCOS. However, there is still lack of comparative evidence that confirms the efficacy of DHA in individuals with PCOS when compared to placebo.

**Methods and analysis:** This is a multicentre randomized, double-blinded, placebo-controlled trial in individuals with PCOS who have irregular menstrual cycles and hyperandrogenism, with or without polycystic ovary morphology. A total of 150 participants will be randomized in a 1:1 ratio to receive oral tablets of 40mg DHA three times per day or placebo for 90 days. The primary outcome is return of regular menstrual cycles within 6 months after start of treatment, with antral follicle count and metabolic profile being secondary outcomes. The primary analysis will follow the intention-to-treat principle.

**Ethics and dissemination:** This study has been approved by the ethics committees of Zhongshan Hospital, Fudan University and all participating centres. All participants will provide written informed consent before randomization. The results will be published in a peer-reviewed journal and will be presented at international meetings.

**Trial registration number:** NCT06842524

**Protocol version:** 1.1 (2025/07/18)

**Strengths and limitations of this study:** This is a multicentre, randomised, double-blinded, placebo-controlled trial.

This study is the first randomized trial to explore the efficacy of DHA in PCOS.

The results may be underpowered to study a smaller than expected effect of DHA.

## INTRODUCTION

Polycystic ovary syndrome (PCOS) is the most common endocrine disorders in individuals of reproductive age, with a prevalence between 10 and 13% ^1^. It encompasses diverse clinical symptoms, including oligoovulation or anovulation, hyperandrogenism, polycystic ovaries, and, in many cases, metabolic disorders ^2^ ^3^. Androgen excess is the main driver of phenotypic features of PCOS, such as follicular dysplasia, impaired ovulation and metabolic dysfunction ^4^. In addition, prenatal androgen exposure induces PCOS-like traits in female offspring, heightening the transgenerational susceptibility to PCOS ^5^. Consequently, controlling androgen excess might be an important pathway in the treatment of PCOS.

Currently, pharmacologic interventions for PCOS are mainly tailored to the management of specific symptoms. Few available drugs effectively target all aspects of PCOS. Combined oral contraceptives (COCs) are recommended for managing hyperandrogenism and/or irregular menstrual cycles in individuals with PCOS. However, COCs do not improve infertility and polycystic ovary morphology. Moreover, COCs may be associated with side effects such as vascular thromboembolism ^6^, which limit their long-term clinical application, particularly for individuals with PCOS with metabolic disorders.

In women, androgen production predominantly occurs in the adrenal glands and ovaries. Within the ovary, synthesis occurs predominantly in theca cells, which are highly responsive to androgenic stimuli like luteinizing hormone (LH) ^7^. Numerous studies have demonstrated an increase of steroidogenic enzymes like CYP11A1 and CYP17A1 in theca cells of PCOS, producing more progesterone, 17a-hydroxyprogesterone, and testosterone ^8–11^. Therefore, inhibiting these enzymes may be effective for controlling hyperandrogenemia and PCOS.

Artemisinin is a well-known antimalarial drug isolated from Artemisia plants ^12^. We have previously demonstrated that artemisinin derivatives can promote energy expenditures and insulin sensitivity by activating thermogenic adipocytes, thereby protecting against diet–induced obesity and metabolic disorders in rodents ^13^ Recently, we demonstrated that artemisinin derivatives could promote the degradation of CYP11A1, thereby inhibiting the synthesis of testosterone in ovarian theca cells ^14^. In rodent models of PCOS, artemisinin derivatives strongly inhibited ovarian androgen synthesis, reduced immature follicles, and improved the estrous cycle. Based on these encouraging findings, we conducted a single arm phase I trial in 19 participants diagnosed with PCOS, with a before and after comparison. Oral intake of dihydroartemisinin (DHA) 40mg tid for 12 weeks normalized menstrual cycles in 63.16% (12/19) of all participants with PCOS, and lowered total testosterone, reduced the antral follicle count ^14^. We now plan to test the efficacy of DHA in a placebo-controlled randomized clinical trial (RCT).

## METHODS AND ANALYSIS

### Study design

This is a multicentre randomised, double-blind, placebo-controlled trial, conducted in four participating centres, namely Zhongshan Hospital, Fudan University (Shanghai, China), The First Affiliated Hospital of Wenzhou Medical University (Wenzhou, China), Women and Children’s Hospital, School of Medicine, Xiamen University (Xiamen, China), and The First Affiliated Hospital of Xiamen University (Xiamen, China).

### Ethics and dissemination

The protocol has been approved by the Ethics Committee in Zhongshan Hospital of Fudan University on February 6, 2025 (Approval No. B2024-417R) and later approved by ethics committees of each participating centre (Approval No. XMYY-2024KY264-03, KY-2025-027-H01, and KY2025-019, respectively). Any change to the study protocol will be documented in a protocol amendment and will be submitted to ethics committee for approval. All participants will provide written informed consent before randomization. Aggregated results will be published in a peer-reviewed journal and will be presented at international meetings. No individual level information will be published or disclosed.

### Participants

We will study individuals with PCOS defined as having irregular menstrual cycles and hyperandrogenism. The detailed inclusion and exclusion criteria for participants are presented below.

Inclusion criteria:

1. Individuals aged 18 - 40 years.
2. 18.5 kg/m^2^ ≤ Body Mass Index (BMI) < 28 kg/m^2^.
3. Negative pregnancy test.
4. No plan for pregnancy in the coming 180 days
5. Two of the Rotterdam criteria for PCOS must be met, namely:

a. Irregular menstrual cycles, which are defined as < 21 or > 35 days or < 8 cycles per year.
b. Hyperandrogenism refers to either hyperandrogenemia or hirsutism. Hyperandrogenemia will be defined as an elevated total testosterone >1.67 nmol/L measured by Elecsys Testosterone II (Roche Diagnostics). Hirsutism is determined by a modified Ferriman-Gallwey Score >4 at screening exam.

Exclusion criteria:

1. Individuals on oral contraceptives. A two-month washout period will be required prior to screening for participants on these agents. A one-month washout will be required for participants on oral cyclic progestins. Individuals on depo-progestins or hormonal implants are ineligible.
2. Individuals with liver disease defined as ALT or AST above normal range of each participating centre, or total bilirubin>30umol/L. Metabolic dysfunction-associated steatotic liver disease (MASLD) with normal ALT and AST can be included.
3. Individuals with anemia (Hemoglobin < 12 g/dL) or neutropenia (neutrocyte <1.8×10^9/^L).
4. Individuals with renal disease defined as serum creatinine> 115umol/L.
5. Individuals diagnosed with other endocrine diseases that are known to cause secondary polycystic ovary morphology, e.g., Cushing’s syndrome, prolactinoma, congenital adrenal hyperplasia (21-hydroxylase deficiency or other enzyme deficiency), hypothyroidism, etc.
6. Individuals diagnosed with Type 1 or Type 2 diabetes.
7. Individuals with known heart disease, like heart failure, atrial fibrillation, coronary heart disease, etc.
8. Individuals with a history of any type of cancer.
9. Individuals taking other medications known to affect reproductive function or metabolism. These medications include GnRH agonists and antagonists, antiandrogens, gonadotropins, GLP-1 receptor agonist, SGLT2i, metformin and thiazolidinediones. The washout period on all these medications will be two months.
10. Individuals who have undergone a bariatric surgery procedure within the past 12 months.

### Informed consent

After screening, eligible individuals will be informed about the study by their physician. If the individual is interested in participating, a dedicated research nurse will discuss the study in detail. Written informed consent is required prior to randomization. All participants are required to take strict contraceptive measures throughout the study period, including male condoms with spermicide (recommended as first choice), uterine caps with spermicide, diaphragm with spermicide, or intrauterine device methods such as copper T-rings. Combined oral contraceptives (COCs) should be avoided throughout the study.

### Withdrawal procedures

Participants will be able to withdraw their consent to take part in the trial at any time. The reason to withdraw will be recorded. If a participant withdraws consent completely, no further data will be collected. If a participant stops medication but agrees to stay in the study, we will continue the follow up, including ultrasound and blood samples. Participants who get pregnant during the study will be required to withdraw from the study.

### Randomization, concealment of allocation and blinding

A computer-generated randomization list in a 1:1 allocation ratio with variable block sizes of 2, 4, or 6 and stratified by the treatment centre will be prepared by an independent statistician who does not participate in the recruitment of participants. Two research staff who are independent of this research team will prepare and label sequentially numbered identical packages of DHA tablets and placebo tablets according to the randomization list.

On the day of initiating treatment, after written informed consent has been provided, participants will be assigned a research number that matches their sequence of enrolment and be provided with packages of medication that match their research number. The allocation sequence and the designated treatment will be completely concealed from the participants, clinicians, investigators, and outcome assessors. Only at the end of the study, when data collection is completed, the allocation sequence will be revealed to the primary investigators. In general, there should be no need to unblind the allocation. If urgent unbinding is necessary, the allocation will be disclosed to the treating physician only.

### Treatment protocols

Participants will be randomised in a 1:1 allocation to DHA tablets 40mg tid for 90 days or Placebo, tid for 90 days. Both DHA tablets and placebo are manufactured by Front Pharmaceutical PLC (Xuancheng, China). They have identical packaging, size, colour and appearance. Treatment will be started between day 2 to day 5 after spontaneous menses or withdrawal bleeding induced by Dydrogesterone (Duphaston) or Estradiol/dydrogesterone (Femoston) administration. For those who have no spontaneous menses throughout the 90 days of treatment of DHA or placebo, Dydrogesterone (Duphaston) or Estradiol/dydrogesterone (Femoston) can be given later to induce withdrawal bleeding. The use of DHA tablets or placebo, along with important precautions, will be explained by a gynecologist, endocrinologist, or a qualified trainee.

### Outcome Measures

#### Primary outcome

The primary outcome measure is the occurrence of a regular menstrual cycle, defined as at least three consecutive spontaneous vaginal bleedings lasting for 2-7 days, with intervals between the start of each cycle of 21 and 35 days (inclusive), during the 26-week period after initiating treatment.

#### Secondary outcomes

1. The presence of a dominant follicle. This will be verified by transvaginal/transanal ultrasound before predicted ovulation, or testing serum progesterone in the predicted mid-luteal phase (A progesterone level>16nmol/L or 5ng/ml is suggestive of ovulation) in those with at least two consecutive spontaneous bleedings. Participants without at least two consecutive spontaneous bleeding will be considered as having no presence of a dominant follicle.
2. The number of bilateral antral follicles. This will be measured by transvaginal ultrasound before and immediately after 90-day treatment. Antral follicles are defined as follicles measuring 2-9 mm in diameter in the ovary.
3. Change in serum AMH. This will be measured before and immediately after 90-day treatment.
4. Change in serum total testosterone, SHBG, and FAI. These will be measured or calculated before and immediately after 90-day treatment. FAI is calculated from measurable values for total testosterone and SHBG, using the following equation: FAI = (Total testosterone in nmol/L / SHBG in nmol/L) × 100.
5. Change in HOMA-IR. Fasting glucose and fasting insulin will be measured and HOMA-IR calculated before and immediately after 90-day treatment. HOMA-IR is calculated using the following equation: HOMA-IR = fasting plasma glucose in mmol/L×fasting insulin in μU/ml/22.5.

### Research visits

All study visits are presented in Table 1. Follow-up procedures and details of each visit are provided in Supplement.

**Table 1.** Follow-up scheme of participants.

|  | Screening | Intervention |  |  |  |  |
| --- | --- | --- | --- | --- | --- | --- |
| Visit | V0 | V1 | V2 | V3 | V4 | V5 |
| Timeline | Within 1 month before baseline | Baseline | 30 ±7 days | 60 ±7 days | 90 ±7 days | 180 ±7 days |
| Screening/Demographic/baseline info |  |  |  |  |  |  |
| Inclusion/Exclusion Criteria | √ |  |  |  |  |  |
| Sign informed consent | √ |  |  |  |  |  |
| Comprehensive history | √ |  |  |  |  |  |
| Record/Dispense medication |  | √ | √ | √ |  |  |
| TSH, creatinine, prolactin | √ |  |  |  |  |  |
| Electrocardiogram (ECG) | √ |  |  |  |  |  |
| Safety measures |  |  |  |  |  |  |
| Pregnancy test | √ | √ | √ | √ | √ | √ |
| Whole blood cell count | √ |  | √ | √ | √ | √ |
| Liver function | √ |  | √ | √ | √ | √ |
| Record adverse events |  |  | √ | √ | √ | √ |
| Efficacy measures |  |  |  |  |  |  |
| Blood Pressure, Heart Rate | √ | √ | √ | √ | √ | √ |
| Body weight, WC, HC, BMI | √ | √ | √ | √ | √ | √ |
| Record menstrual details | √ | √ | √ | √ | √ | √ |
| Ultrasound | √ | √* |  |  | √ | √ |
| T, FSH | √ | √* |  |  | √ | √ |
| AMH, SHBG, FAI, DHEA-S |  | √* |  |  | √ | √ |
| LH, E2, P |  | √* |  |  | √ | √ |
| Fasting glucose & insulin |  | √* |  |  | √ | √ |
| HbA1c |  | √* |  |  | √ | √ |
| TG, TC, LDL, HDL |  | √* |  |  | √ | √ |
| Quality of life questionnaire |  | √ |  |  | √ | √ |

\* Results within four weeks before medication are acceptable.

### Data collection

All data including basic characteristics, clinical and laboratory outcomes will be recorded into participants’ electronic medical records as part of routine clinical care. Data from the included participants will be compiled utilizing custom electronic case report forms (eCRFs) and inputted into the electronic database by the research staff of each participating centre. This data will be subjected to several validation checks to confirm the accuracy and comprehensiveness of the captured data.

### Sample size calculation

In a pilot study at Zhongshan Hospital Fudan University, the rate of menstrual cycle normalization in PCOS individuals using DHA was 36.7% (11/30). The occurrence of a regular menstrual cycle in the placebo group is estimated to be no more than 10%. An increase of 25% in normalization rate in DHA-treated group vs placebo is considered to be highly clinically relevant. In order to detect or refute a 25% increase (power 90%, two-sided significance level of 0.05), each group should have at least 57 participants. Considering a possible 25% loss to follow up rate, we plan to recruit 75 participants in each group (150 participants in total).

### Statistical analysis

The primary analysis will be conducted on an intention-to-treat basis. Per-protocol analyses will be conducted but these will be considered exploratory only. The per-protocol population will exclude those who did not receive the intervention/placebo or those with poor adherence (intake less than 75% of medication within 90 days). Baseline data will be presented using descriptive statistics (mean and standard deviation for normally distributed variables, or median and interquartile range for skewed variables). Categorical data will be presented as numbers (%). The analysis of all outcomes will adjust for the study centre. For binary outcomes, we will estimate adjusted risk ratios (aRRs) with 95% CIs using a generalized linear model, with Poisson log link function and robust variance estimate. For continuous outcomes or difference in differences outcomes, generalized linear models with negative binomial distribution or binomial distribution will be used. Missing data for the primary outcome will be handled with sensitivity analysis using the worst-case scenario and multiple imputation. Details of data analysis will be described in a separate statistical analysis plan, which will be finalized before collecting the last participant’s data.

### Interim analysis & Data Safety Monitoring Committee

An interim analysis will be carried out after enrollment of 100 participants. An independent statistician, blinded to treatment allocation, will prepare a report focusing on safety and the primary outcome. This report will be provided exclusively to the Data Safety Monitoring Committee (DSMC). An interim analysis of the primary outcome will be conducted using a two-sided significance test, applying the Haybittle-Peto boundary for early efficacy monitoring. To preserve an overall Type I error rate of 5%, interim findings will require a p-value less than 0.001 (Z = 3.29) to be considered statistically significant.

The independent DSMC will review the progress of the trial regularly and provide advice on the conduct of the trial to the Trial Steering Committee (TSC), who will consider and act, as appropriate, and ultimately carries the responsibility for deciding whether a trial needs to be stopped on grounds of efficacy and safety. Also, the DSMC will monitor serious adverse events that have occurred. The DSMC consists of an independent chair specialized in PCOS, a reproductive endocrinologist and a biostatistician.

### Safety reporting

The investigator will inform participants, the DSMC and the reviewing accredited medical research ethics committee if any safety concerns occur, on the basis of which it appears that the disadvantages of participation may be significantly greater than was foreseen in the research proposal.

### Adverse and serious adverse events

All observed or volunteered adverse events, regardless of treatment group or suspected causal relationship to intervention, will be recorded. Adverse events are defined as any undesirable experience occurring to a participant during the trial, whether or not considered related to the intervention. All adverse events reported spontaneously by the participant or observed by the investigator or their staff will be recorded. A serious adverse event (SAE) is any untoward medical occurrence or effect, at any dose, that results in death; is life threatening (at the time of the event); requires hospitalization or prolongation of existing inpatients’ hospitalization; results in persistent or significant disability or incapacity; is a new event of the trial likely to affect the safety of the participants, such as an unexpected outcome of an adverse reaction. All SAEs will be reported to the accredited medical research ethics committee that approved the protocol. Participants will be covered by clinical trial insurance in the event they suffer injury or harm as a result of trial participation.

## DISCUSSION

Artemisinin derivatives have shown great promise in various applications with minimum adverse effects, such as treating malaria, cold, diarrhea, lupus erythematosus, and cancer ^12^. Nevertheless, a possible effect of artemisinin derivatives on PCOS has not been studied in a clinical trial. New data suggest that dysregulated metabolic pathways are involved in PCOS development, including impaired thermogenic adipose tissue ^15^, the microbiota-gut ovary axis ^16^, and systemic insulin resistance ^17^.

Our previous findings showed that artemisinin derivatives promoted metabolic homeostasis and protected against obesity ^13^, which prompted us to investigate a potential role of artemisinins in PCOS. We observed beneficial effects of artemisinin derivatives in alleviating typical phenotypes of PCOS in rodent models, and further confirmed the efficacy of DHA in ameliorating hyperandrogenism, irregular menstrual cycles, and polycystic ovary morphology in individuals with PCOS ^14^. However, we did not see a significant metabolic impact of artemisinin derivatives in PCOS rodents, implying that the therapeutic effect might not rely on improving systemic metabolism.

This placebo controlled randomized clinical trial will address the potential feasibility of using DHA as a therapeutic agent for PCOS. Both phenotype-related and metabolic outcomes are set to be evaluated in this trial. The results will have a potential impact on the standard of care for participants diagnosed with PCOS.

## Supporting information

Supplemental Appendix to Study Protocol

## Data Availability

Data supporting the findings will be available upon reasonable request to the corresponding author after the completion of the study.

## Trial status

The trial recruitment has already started in April 2025 with an expected duration of 18 months for recruitment. It is expected to recruit the last participant in September 2026.

## Acknowledgements

We thank the DSMC for providing inputs on the trial management. We also thank all participants who will be involved in this trial.

## Members of DSMC

Dr. Robert Norman from The University of Adelaide (Chair of DSMC), Dr. Tony Zhang from The University of New South Wales, and Dr. Miaoxin Chen from Tongji University.

## Contributors

Study concept and design: Jingjing Jiang, Liangshan Mu, Xi Dong, and Ben Willem Mol. Drafting of the manuscript: Jingjing Jiang, Liangshan Mu, and Yanting Shen. Critical revision of the manuscript for important intellectual content: Jingjing Jiang and Liangshan Mu. Study supervision: Ben Willem Mol, Wentao Li, Xi Dong, and Xiaoying Li. Recruitment: Yanting Shen, Liangshan Mu, Jingjing Jiang, Guiquan Wang, Changqin Liu, Haiyan Yang. Statistical analysis: Wentao Li. Critical discussion: Ben Willem Mol, Wentao Li, Liangshan Mu, and Jingjing Jiang.

## Competing interests

BWM is supported by a NHMRC Investigator grant (GNT1176437). BWM reports consultancy, travel support and research funding from Merck and consultancy for Ferring, Organon, UNILAB and Norgine. WL declared research grant support from the Australian National Health and Medical Research Council, the Medical Research Future Fund, and the Ramaciotti Foundations for works unrelated to the current topic. Other authors report no conflict of interest.

All the dihydroartemisinin tablets and placebo are manufactured and donated by Front Pharmaceutical PLC. Front Pharmaceutical PLC has no role in the design, participant enrollment, treatment, follow-up, data analysis, and publication of any results.

## Patient and public involvement

Patients and/or the public were not involved in the design, or conduct, or reporting, or dissemination plans of this research.

## Patient consent for publication

Not required.

## Provenance and peer review

Not commissioned; externally peer reviewed.

## Open access

This is an open access article distributed in accordance with the Creative Commons Attribution Non Commercial (CC BY-NC 4.0) license, which permits others to distribute, remix, adapt, build upon this work non-commercially, and license their derivative works on different terms, provided the original work is properly cited, appropriate credit is given, any changes made indicated, and the use is non--commercial. See: http://creativecommons.org/licenses/by-nc/4.0/.

## Data sharing plan

Following completion of the trial and publication of the results by the trial team, deidentified participant data and analysis code will be made available upon reasonable request through the corresponding authors. Requests must be accompanied by a scientifically and ethically sound proposal, which will be reviewed by the trial team prior to approval. The statistical analysis plan will be included as a supplementary file in the trial publication.

## Confidentiality Statement

Personal information of potential and enrolled participants will be collected, stored, and managed in accordance with China’s privacy laws to ensure confidentiality. Each participant will be assigned a unique study ID, and identifying details will be stored separately from study data in secure, access-controlled systems. Only authorised personnel will have access to identifiable information. Data shared for analysis or reporting will be de-identified. Confidentiality will be maintained before, during, and after the trial.

