## Supplemental Appendix to Study Protocol for "Dihydroartemisinin for Treatment of Polycystic Ovary Syndrome: study protocol for a multi-centre placebo-controlled Randomized Clinical Trial"

**Follow-up procedures**

Screening Visit (Visit 0: Within one month before Baseline)

(1) Take a comprehensive physical examination (weight, height, waist circumference, hip circumference, BMI, blood pressure); take a comprehensive history: past medical history (what kind of disease was diagnosed and what kind of treatment was obtained during the past years), family history, reproductive history (including pregnancy time, abortion time and so on).
(2) Document detailed menstrual history of the past six months, and all the medications used to treat PCOS, including but not limited to: COCs, Progestins, metformin, TZDs, GLP-1RA, etc.
(3) FSH, total testosterone, β-hCG, TSH.
(4) Perform transvaginal ultrasound to document antral follicle counts and ovary volume.
(5) Safety parameters: liver and renal function, complete blood count, ECG.

Visit 1 (V1: Baseline)

(1) LH, FSH, estrogen, total testosterone, progesterone, SHBG, DHEAS, AMH, β-hCG, fasting glucose, fasting insulin, HbA1c, lipid profiles (Results except β-hCG within four weeks before medication are acceptable).
(2) Perform transvaginal ultrasound to document antrol follicle counts and ovary volume (Results within four weeks before medication are acceptable).
(3) Safety parameters: liver and renal function, complete blood count, ECG (Results within four weeks before medication are acceptable).
(4) Quality of Life measured by Polycystic Ovary Syndrome Health-Related Quality of Life Questionnaire [PCOSQ] score, and 36-Item Short Form Health Survey [SF-36].
(5) Ensure patient agree to sign informed consent.
(6) Randomization.
(7) Take blood sample for repository in central lab.
(8) Dispense study medication.

Visit 2 and 3 (30 days and 60 days after starting medication)

(1) Record weight, waist circumference, hip circumference, BMI, blood pressure.
(2) Document menstrual event in detail if there is any.
(3) Liver function, complete blood count, and β-hCG.
(4) Record adverse events and concomitant medications if there is any.
(5) Dispense study medication.

Note: 1. If there is an elevation of ALT or AST above 3 times upper limit, DHA should be stopped. If the increase of ALT or AST is below 3 times upper limit. DHA can be maintained, and drugs like silymarin, Glycyrrhizin, etc can be added. 2. If there is a drop of neutrophils below $1.4\times10^9$/L, DHA should be stopped. If neutrophil count is lower than $1.8\times10^9$/L, but still above $1.4\times10^9$/L, DHA can be maintained, and drugs like leucogen, etc can be added. 3. If there is a drop of hemoglobin below 11g/dL, DHA should be stopped.

Visit 4 (90 days after starting medication)

(1) Record weight, waist circumference, hip circumference, BMI, blood pressure.
(2) Document menstrual events in detail.
(3) LH, FSH, estrogen, total testosterone, progesterone, SHBG, DHEAS, AMH, β-hCG, fasting glucose, fasting insulin, HbA1c, lipid profiles.
(4) Perform transvaginal ultrasound to document antral follicle counts and ovary volume.
(5) Safety parameters: liver function, complete blood count.
(6) Quality of Life measured by Polycystic Ovary Syndrome Health-Related Quality of Life Questionnaire [PCOSQ] score, and 36-Item Short Form Health Survey [SF-36].
(7) Record adverse events and concomitant medications if there is any.
(8) Take blood sample for repository in central lab.

Visit 5 (180 days after starting medication)

(1) Record weight, waist circumference, hip circumference, BMI, blood pressure.

(2) Document menstrual events in detail.

(3) LH, FSH, estrogen, total testosterone, progesterone, SHBG, DHEAS, AMH, β-hCG, fasting glucose, fasting insulin, HbA1c, lipid profiles.

(4) Perform transvaginal ultrasound to document antrol follicle counts and ovary volume.

(5) Safety parameters: liver function, complete blood count.

(6) Quality of Life measured by Polycystic Ovary Syndrome Health-Related Quality of Life Questionnaire [PCOSQ] score, and 36-Item Short Form Health Survey [SF-36].

(7) Record adverse events and concomitant medications if there is any.

Note: Patients are followed up monthly by phone call or wechat. If there are at least two consecutive menstrual events, with intervals between 21 and 35 days, patients are required to come back at least once for a transvaginal ultrasound to confirm the presence of a dominant follicle before predicted ovulation, or testing serum progesterone in the predicted midluteal phase (A progesterone level>16nmol/L or 5ng/ml is suggestive of ovulation).

**Procedures for monitoring adherence**

Research staff at each study site will review dosing information with each patient on scheduled clinic visits, providing detailed instructions regarding dose frequency and the number of tablets to be taken for each dose. Patients will be instructed to keep all unused containers (empty, partially used, and/or unopened) for accountability at the next scheduled clinic visits. A compliance check and tablet count will be performed by research staff during clinic visits and the results will be recorded.

**Strategies for achieving adequate participant enrollment**

Participants are recruited from Departments of Endocrinology, Gynecology, and Reproductive Medicine at each participating centre. Potential candidates in the outpatient clinic will be screened and eligible individuals will be recruited by clinical staff who are trained and familiar with the study protocol. Recruitment advertisement will be posted in the outpatient clinic as well as in social media to enhance the visibility of the study among potential candidates.
